# Age-dependent Dynamics of the Electrocardiographic Parameters in Cardiovascular Disease-Free Children

**DOI:** 10.64898/2026.02.07.26345587

**Authors:** Kazi T. Haq, Kathleen McLean, Charles I. Berul, Nikki Gillum Posnack

## Abstract

**Background:** Normative pediatric electrocardiographic (ECG) parameters are standardized, but lack temporal resolution for neonates and infants. These values are clinically important, as they support the diagnosis, risk stratification, and management of cardiovascular diseases (CVD).

**Methods:** Five ECG parameters (heart rate (HR), QRS, PR, QT, QTc intervals) were retrospectively analyzed from 7,346 recordings from 6,967 patients at a large pediatric hospital. Patients were only included if their ECG was adjudicated as normal by a pediatric cardiologist. Patients were assigned to 45 age groups: neonates (1-35 days, 35 groups), infants (2-6 months, 5 groups), and young children (1-5 years, 5 groups). Sensitivity analysis ranked ECG parameters to determine those most affected by age. Z-scores were used to quantify deviations in developmental ECG parameter trajectories in CVD-free patients compared with unrepaired tetralogy of Fallot (TOF, n=305).

**Results:** Developmental shifts in ECGs were observed for all patients, irrespective of whether intensive care unit or CVD patients were included in the analysis. All five ECG parameters differed significantly between early (1-8 days) and late neonates (9-35 days). Sensitivity analysis revealed rapid ECG parameter adaptations during the neonatal stage, with slower changes during infancy and early childhood. Unrepaired TOF patients had significantly different HR, PR, and QRS values in the late neonatal group compared with CVD-free children. Z-scores revealed disease-specific deviations (≥ 2 SD of baseline), including outlier QTc values in 32.7% and 24.3% of early and late neonatal TOF patients, respectively.

**Conclusion:** This study defined the values of five key ECG parameters, with enhanced age-specific resolution in neonates, infants, and children. Neonatal age emerged as the most dynamic stage for ECG parameter changes. This study demonstrated that high temporal resolution of age-specific ECG parameters can identify abnormal deviations, which can provide valuable insight for the diagnosis and management of CVD in early life.

**CLINICAL PERSPECTIVE:** *What is known:*

- Normative pediatric ECG values derived from healthy children have been reported; however, available reference datasets are often limited by broad age stratification, particularly during the neonatal and infant periods.

*What the study adds:*

- High-temporal-resolution reference values are reported for five clinically relevant ECG parameters, with day-level resolution in neonates and month-level resolution in infants.
- Retrospective ECG data from a CVD-free pediatric cohort closely align with values reported in healthy pediatric populations, supporting their use as surrogate baseline reference data.
- Postnatal ECG developmental trajectories are preserved in CVD-free pediatric patients but deviate in disease states such as unrepaired tetralogy of Fallot, highlighting the potential clinical utility of these reference trajectories for identifying disease-related alterations.

## INTRODUCTION

The cardiovascular system undergoes profound changes during the perinatal transition from intra-to extrauterine life^1,2^. These physiological adaptations are most evident during the postnatal phase, as it includes an abrupt shift from fetal to pulmonary/systemic circulation. Initially, the right ventricle is the dominant chamber due to fetal circulatory patterns; yet, within weeks after birth, the left ventricle increases in size and gains dominance as fetal shunts close, pulmonary resistance decreases, and vascular resistance increases. Postnatal remodeling alters not only the size and mass of the cardiac chambers, but also induces developmental shifts in autonomic regulation and cardiac conduction. These developmental adaptations are reflected by measurable changes in the cardiac electrocardiographic (ECG) parameters, denoted by shifts in heart rate, axis deviation, and interval durations^3,4^. Postnatal electrical remodeling occurs over a compressed timeline, as ECG parameters evolve on a week-by-week basis within the first few months of life^5^. After infancy, additional changes in ECG morphology occur gradually throughout childhood and adolescence. Accordingly, non-invasive ECG recordings are a critical tool for identifying electrical and/or structural abnormalities – but, this requires interpretation within the context of a patient’s developmental stage.

Normative pediatric ECG values derived from healthy children have been reported previously ^5–10;^ however, these reference datasets are often limited by broad age stratification, particularly during the neonatal, infant, and early childhood periods. Such coarse grouping can obscure subtle yet clinically meaningful age-related transitions in cardiac electrophysiological maturation. Moreover, it remains uncertain whether ECG reference standards established in healthy volunteer populations are directly applicable to children who undergo clinically indicated ECG recordings in a hospital setting, even in the absence of diagnosed cardiovascular disease (CVD). Importantly, acquisition of high–temporal resolution ECG data (e.g., day-by-day measurements in neonates) from truly healthy individuals is inherently challenging due to ethical considerations and practical barriers to enrollment at early developmental stages. To address these limitations, we leveraged a CVD–free pediatric cohort receiving clinical care to establish age-resolved baseline ECG metrics across childhood. In the absence of high–temporal resolution, age-specific ECG reference standards, it may be challenging to distinguish normal developmental physiology from pathophysiology (e.g., congenital heart disease, cardiomyopathy, or arrhythmia), particularly when available datasets are derived from narrowly defined study populations. This limitation is especially consequential in emergency departments and intensive care units (ICUs), where pediatric ECGs are frequently used to evaluate acute presentations (e.g., chest pain, seizure or syncope, dysrhythmia, toxicologic screening^11,12^) and timely clinical decision-making is critical. Establishing high-resolution, age-specific ECG reference standards has the potential to improve the sensitivity and specificity of ECG-based screening, facilitate earlier and more accurate clinical interventions, and reduce false-positive findings or unnecessary treatment. Moreover, robust pediatric ECG parameter reference standards will become increasingly important as automated analysis tools and machine learning–based approaches are integrated into clinical workflows for disease detection and risk stratification^13–16^.

In prior work, we showed that age is a key demographic factor that strongly influences ECG metrics in both humans and animal subjects^17–21^. Based on these findings, we hypothesized that clinically important ECG parameters vary substantially across days in neonates, months in infants, and years in young children, reflecting postnatal cardiac remodeling. The goals of this study were to establish high–temporal resolution reference ranges for five clinically relevant ECG parameters—heart rate (HR), PR interval, QRS duration, QT interval, and heart rate–corrected QT (QTc)—and to determine whether a CVD–free pediatric cohort preserves age-dependent transitional features previously observed in healthy populations.

To achieve these goals, we retrospectively analyzed ECG data from 6,967 pediatric patients with normal ECG recordings, as determined by pediatric cardiologist interpretation, stratified into 45 age groups. We first evaluated whether developmental trends in ECG parameters were consistent across clinically distinct subgroups of patients with normal ECGs. We then compared ECG parameters derived from our CVD-free patient cohort with two previously published datasets obtained from healthy children^5,8^. Finally, as a case study, we examined patients with unrepaired tetralogy of Fallot (TOF) to demonstrate deviations in age-dependent ECG trajectories relative to age-matched CVD-free patients.

## METHODS

### Study population and exclusion criteria

In this retrospective IRB approved study (#12146), ECG datasets were collected from 6,967 patients who visited Children’s National Hospital between 1994 and 2024. Standard 12-lead ECGs were recorded using the MAC 5500 ECG system and collected from the MUSE v9 Cardiology Information System (GE Medical Systems). ECG recordings with significant noise, missing leads, or signal quality issues were excluded. Individuals were only included if they had a normal ECG, as classified by the consulting physician. ECG recordings were binned into 45 age groups with resolution in days for neonates, months for infants, and years for children. Patients were divided into three cohorts 1) all patients (n=6,967; **Table 1**), 2) excluding ICU patients (n=5,007), 3) CVD-free cohort (excluding both ICU and patients with CVD, n=2,449; **Supplemental Table 1-2**). For statistical analysis between ages, patients were binned into four larger age groups: early neonate (1-8 days), late neonate (9-35 days), infant (2-6 months), and early childhood (1-5 years). To assess disease-specific deviations, ECGs were analyzed from unrepaired (preoperative) TOF patients (n=305) without bundle branch block. For comparisons with published datasets in healthy individuals, we utilized the Brantincsak, et al. study^5^ (healthy children and adults, ages 0-39 years) and the NIH/NHLBI Pediatric Heart Network (PHN) Echo-Z/Normal ECG dataset (healthy children, ages 0-18 years) ^8^. For the latter, data were downloaded from www.pediatricnetwork.org on 02/11/2025.

**Table 1.** Mean ECG values for all patients (n=6,967).

| Age<br>(days) | HR<br>(mean) | HR<br>(SEM) | PR<br>(mean) | PR<br>(SEM) | QRS<br>(mean) | QRS<br>(SEM) | QT<br>(mean) | QT<br>(SEM) | QTc<br>(mean) | QTc<br>(SEM) |
| --- | --- | --- | --- | --- | --- | --- | --- | --- | --- | --- |
| *Early Neonate* |  |  |  |  |  |  |  |  |  |  |
| 1 | 123.52 | 3.00 | 117.44 | 2.98 | 62.76 | 2.62 | 302.56 | 4.84 | 423.49 | 3.96 |
| 2 | 130.53 | 1.62 | 113.73 | 2.14 | 57.08 | 1.24 | 288.54 | 2.58 | 423.42 | 2.61 |
| 3 | 132.12 | 1.68 | 109.03 | 2.65 | 55.83 | 1.61 | 287.76 | 2.96 | 422.37 | 3.04 |
| 4 | 129.28 | 1.79 | 108.77 | 2.06 | 55.79 | 1.31 | 286.67 | 2.62 | 416.50 | 2.31 |
| 5 | 133.12 | 1.70 | 106.99 | 1.68 | 53.46 | 1.25 | 283.31 | 2.35 | 418.66 | 2.53 |
| 6 | 133.96 | 2.05 | 103.35 | 1.97 | 56.40 | 1.61 | 281.06 | 3.46 | 413.75 | 2.89 |
| 7 | 140.90 | 1.35 | 107.58 | 1.49 | 55.46 | 1.00 | 275.95 | 1.87 | 420.10 | 2.25 |
| 8 | 143.03 | 1.61 | 102.61 | 1.44 | 53.94 | 1.01 | 268.69 | 2.28 | 411.12 | 2.34 |
| *Late Neonate* |  |  |  |  |  |  |  |  |  |  |
| 9 | 146.97 | 1.53 | 105.31 | 1.37 | 53.35 | 0.90 | 262.72 | 1.72 | 408.93 | 1.83 |
| 10 | 149.26 | 1.29 | 104.80 | 1.42 | 54.54 | 0.94 | 263.78 | 1.78 | 413.51 | 2.22 |
| 11 | 148.99 | 1.39 | 105.56 | 1.45 | 56.00 | 1.23 | 266.52 | 1.78 | 417.67 | 2.03 |
| 12 | 148.63 | 1.35 | 106.46 | 1.40 | 55.74 | 1.27 | 267.94 | 1.72 | 417.75 | 1.81 |
| 13 | 151.48 | 1.39 | 105.99 | 1.27 | 54.88 | 1.00 | 261.89 | 1.60 | 413.73 | 1.58 |
| 14 | 145.96 | 1.31 | 104.55 | 1.37 | 55.81 | 0.93 | 268.91 | 1.69 | 416.43 | 1.73 |
| 15 | 151.24 | 1.23 | 105.25 | 1.12 | 55.56 | 0.98 | 263.31 | 1.62 | 415.62 | 1.74 |
| 16 | 150.60 | 1.23 | 102.87 | 1.05 | 54.51 | 0.73 | 265.04 | 1.56 | 418.96 | 1.94 |
| 17 | 150.30 | 1.28 | 104.92 | 1.27 | 55.57 | 0.75 | 266.46 | 1.66 | 419.58 | 1.89 |
| 18 | 153.27 | 1.40 | 106.91 | 2.60 | 56.90 | 1.11 | 262.15 | 2.26 | 414.62 | 3.34 |
| 19 | 151.76 | 1.23 | 104.25 | 1.16 | 54.50 | 1.08 | 263.40 | 1.51 | 416.96 | 1.58 |
| 20 | 150.04 | 1.26 | 103.27 | 1.09 | 54.44 | 0.72 | 266.82 | 1.92 | 418.09 | 1.78 |
| 21 | 152.50 | 0.94 | 103.91 | 0.98 | 54.62 | 0.53 | 264.58 | 1.25 | 419.04 | 1.42 |
| 22 | 154.99 | 1.04 | 104.08 | 1.11 | 55.18 | 0.75 | 260.28 | 1.35 | 416.18 | 1.61 |
| 23 | 155.14 | 1.10 | 102.90 | 1.12 | 53.40 | 0.71 | 260.93 | 1.47 | 417.56 | 1.48 |
| 24 | 152.30 | 1.17 | 104.93 | 1.20 | 54.77 | 0.90 | 263.86 | 1.58 | 419.23 | 1.72 |
| 25 | 153.36 | 1.38 | 104.90 | 1.18 | 54.97 | 0.88 | 262.41 | 1.54 | 417.24 | 1.50 |
| 26 | 152.67 | 1.25 | 103.15 | 1.15 | 52.79 | 0.85 | 265.77 | 1.57 | 422.63 | 1.74 |
| 27 | 153.19 | 1.20 | 106.52 | 1.06 | 54.78 | 0.87 | 265.70 | 1.45 | 422.62 | 1.62 |
| 28 | 151.76 | 1.19 | 103.37 | 1.07 | 55.91 | 0.84 | 267.30 | 1.63 | 422.94 | 1.86 |
| 29 | 153.91 | 1.00 | 103.10 | 1.04 | 53.70 | 0.68 | 262.83 | 1.74 | 420.31 | 1.28 |
| 30 | 154.26 | 1.17 | 101.70 | 0.92 | 55.13 | 0.84 | 265.51 | 1.51 | 422.48 | 1.49 |
| 31 | 154.58 | 1.37 | 101.79 | 1.25 | 52.86 | 0.82 | 262.75 | 1.91 | 419.61 | 2.44 |
| 32 | 156.41 | 1.22 | 104.86 | 1.67 | 54.19 | 1.01 | 262.64 | 1.57 | 422.11 | 1.61 |
| 33 | 154.44 | 1.39 | 103.62 | 1.85 | 54.31 | 1.33 | 262.79 | 1.61 | 420.29 | 1.99 |
| 34 | 152.72 | 1.46 | 104.14 | 1.45 | 55.60 | 0.77 | 263.77 | 1.69 | 418.84 | 1.84 |
| 35 | 154.70 | 1.18 | 102.97 | 1.61 | 54.72 | 1.04 | 263.34 | 1.85 | 419.97 | 1.87 |
| *Infant* |  |  |  |  |  |  |  |  |  |  |
| 60 | 147.36 | 0.68 | 100.90 | 0.65 | 57.75 | 0.43 | 270.22 | 0.84 | 421.15 | 0.81 |
| 90 | 140.69 | 1.17 | 100.85 | 0.93 | 62.27 | 1.35 | 279.10 | 1.42 | 424.46 | 1.34 |
| 120 | 138.77 | 1.05 | 103.59 | 0.95 | 61.56 | 0.59 | 281.34 | 1.40 | 425.33 | 1.41 |
| 150 | 137.27 | 0.98 | 105.99 | 0.84 | 63.12 | 0.61 | 281.03 | 1.21 | 422.80 | 1.21 |
| 180 | 132.59 | 0.86 | 106.32 | 0.88 | 62.35 | 0.70 | 285.91 | 1.26 | 421.70 | 1.24 |
| *Early Childhood* |  |  |  |  |  |  |  |  |  |  |
| 365 | 118.61 | 1.18 | 116.25 | 0.85 | 65.54 | 0.55 | 296.16 | 1.09 | 415.70 | 1.03 |
| 730 | 114.20 | 0.98 | 119.87 | 0.90 | 66.59 | 0.48 | 299.21 | 1.24 | 407.89 | 0.86 |
| 1095 | 106.24 | 0.97 | 124.20 | 1.01 | 70.56 | 0.49 | 319.54 | 1.49 | 420.20 | 1.14 |
| 1460 | 103.26 | 1.00 | 122.68 | 0.98 | 73.47 | 0.62 | 324.99 | 1.36 | 420.50 | 1.14 |
| 1825 | 96.92 | 1.07 | 127.21 | 1.04 | 74.34 | 0.64 | 333.08 | 1.65 | 418.55 | 1.17 |

### Data representation and statistical analysis

Data are reported as mean ± standard error mean (SEM), unless otherwise indicated. Group comparisons were analyzed using either a T-test or ordinary one-way ANOVA with Holm-Sidak multiple comparisons testing. Statistical significance was defined as a corrected p-value < 0.05, which is denoted in each figure by an asterisk.

Longitudinal vectorcardiographic loops of a patient (5 months-27 years) were constructed from 12-lead ECG using previously described methods ^22^. Briefly, standard 12 lead ECG signals were preprocessed using noise filtering and baseline wander removing. After detecting R peaks and aligning individual beats, outlier beats were excluded and a median beat was computed for all 12 leads. The median 12-lead beats were then transformed into orthogonal X, Y, and Z signals using the Kors regression coefficients. These orthogonal signals were subsequently rotated into an anatomical reference frame to reconstruct the 3-dimensional QRS-T vector loop. Standard frontal, horizontal, and sagittal projections were generated from the same transformed trajectories.

To quantify the developmental sensitivity of ECG parameters within each age bin, we defined a normalized sensitivity parameter (α) that captures the relative magnitude of physiological change per unit time. Specifically, sensitivity was calculated as the trimmed 90% range of an ECG metric, normalized by both the duration of the age interval and the median value of the metric:

Sensitivity parameter were defined as:

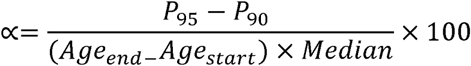

where, *P*_95_ - *P*_90_ =the trimmed 90% range of an ECG metric

*Age_end-_Age_start_* =the number of days in the age bin

This formulation expresses sensitivity as a percentage change per day (%/day), allowing direct comparison of developmental dynamics across ECG parameters with different physical units and scales. By using a trimmed range rather than extrema, the metric is robust to outliers and measurement noise inherent in retrospective clinical ECG data.

To quantify disease-related deviations in ECG parameters, Z-scores were calculated relative to baseline values derived from the CVD-free cohort. For each ECG parameter and age group, the mean (μ) and standard deviation (σ) were computed using the CVD-free cohort and used as reference values. Z-scores for individual ECG measurements were then calculated as:

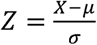

where *X* represents the observed ECG parameter value in an individual patient. Z-scores were subsequently used to assess deviations in the unrepaired TOF cohort relative to age-matched CVD-free baseline values, with abnormal values defined as deviations ≥ 2 standard deviations from the reference mean.

## RESULTS

### Dynamic adaptations in ECG parameters occur early in postnatal life

Prior studies reported age-dependent ECG metric changes in healthy children^5–10;^however, these studies combined individuals into relatively broad age groups (e.g., <1month^8^ or 1-4 weeks^5^). Notably, we observed compelling day-to-day variations in ECG parameters within the first month of postnatal life. Across the entire cohort of pediatric patients, heart rate increased 15.8% within the first eight days (1 day: 123.5±3 bpm, 8 days: 143.0±1.6 bpm) and 25.3% within the 35 days (154.7±1.2 bpm; **Figure 1A**). Other ECG parameters changed rapidly within the first week, then reached a plateau in older neonates. The latter included shortening of the PR interval by 12.6% (1 day: 117.4±3, 8 days: 102.6±1.4 ms) and 12.3% (35 days: 103±1.6 ms; **Figure 1B**), shortening of the QRS complex by 14.2% (1 day: 62.7±2.6, 8 days: 53.9±1 ms) and 12.9% (35 days: 54.7±1 ms; **Figure 1C**), and shortening of the QT interval by 11.2% (1 day: 302.6±4.8, 8 days: 268.7±2.3 ms) and 13% (35 days: 263.3±1.8 ms; **Figure 1D**). The QTc interval stayed relatively stable, shortening by 2.9% (1 day: 423.5±4, 8 days: 411.1±2.3 ms) and 0.8% (35 days: 420±1.9 ms; **Figure 1E**). It is important to note that we employed the Bazett formula for QT correction because of its broad clinical use; however, this formula can result in overcorrection at faster heart rates in pediatric subjects^17^.

**Figure 1.**
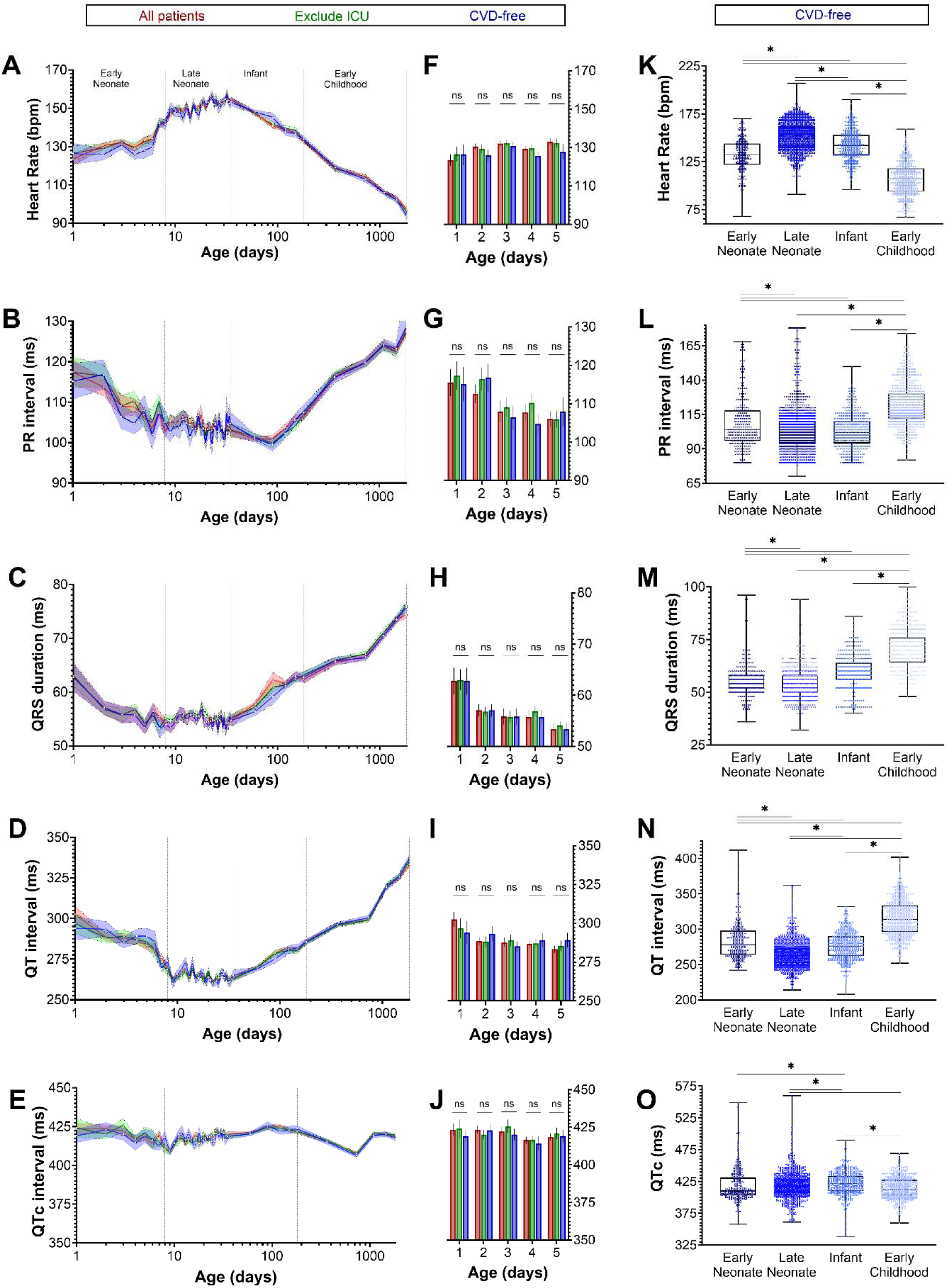
Age-dependent baseline ECG characteristics in the pediatric population. Derived ECG values for heart rate, PR interval, QRS duration, QT interval, and QTc interval are shown. **A-E)** Mean ECG values for 45 age groups including all patients (red), excluding ICU patients (green), and CVD-free (blue). Mean value indicated by solid line; standard error of the mean indicated by color bands. **F-J)** No significant differences were observed between patient groups, even during early neonatal development (1-5 days). Unpaired two-tailed T-test, ns= not significant. **K-O)** Individual datapoints shown for patients in the CVD-free cohort, binned into four age groups: early neonate (1-8 days), late neonate (9-35 days), infant (2-6 months), and early childhood (1-5 years). Boxplots represent the median, lower and upper quartiles. One-way ANOVA with Hold-Sidak multiple comparisons testing, *p<0.05.

After the neonatal period, subsequent ECG parameter changes tended to shift in the opposite direction. For example, heart rate slowed by 14.3% at 6 months (35 days: 154.7±1.2, 6 months: 132.6±0.9 bpm) and 37.4% at 5 years (96.9±1.1 bpm; **Figure 1A**). During this timeframe the PR interval progressively lengthened by 3.2% (35 days: 103±1.6, 6 months: 106.3±0.9 ms) and 23.5% (5 years: 127.2±1 ms; **Figure 1B**), QRS lengthened by 14.1% at 6 months (35 days: 54.7±1, 6 months: 62.4±0.7 ms) and 35.8% (5 years: 74.3±0.6 ms; **Figure 1C**), and QT lengthened by 8.9% (35 days: 263.3±1.8, 6 months: 285.9±1.3 ms; **Figure 1D**) and 26.5% (5 years: 333.1±1.7 ms). The QTc interval stayed relatively stable, with a modest 0.4% (35 days: 420±1.9, 6 months: 421.7±1.2 ms) and 0.3% change (6 years: 418.6±1.2 ms; **Figure 1E**).

### Age-dependent shifts in ECG parameters are observed across different patient cohorts

Initially, ECG parameters were analyzed and averaged across a clinically diverse pediatric cohort encompassing both inpatient and outpatient cardiology assessments, although all patients had ECG recordings classified as normal by a cardiologist. However, medical interventions and medications can significantly alter ECG parameters, including those commonly administered to neonatal and pediatric ICU patients (e.g., analgesics, sedatives, antibiotics) and/or patients with CVD (e.g., inotropes, vasopressors, antiarrhythmics)^23,24^. Accordingly, we assessed whether these age-dependent adaptations persisted across three patient cohorts: 1) all patients, 2) ICU patients excluded, and 3) CVD-free (ICU and CVD patients excluded). We observed very similar developmental trends in all three patient cohorts across all ECG parameters (**Figure 1A-E**). While we observed slightly larger deviations <u>within</u> each cohort in the early neonatal period (e.g., 1-5 days), there was no significant difference <u>between</u> the three cohorts during this early stage of development (**Figure 1F-J**). With advancing age, the developmental trajectories were nearly identical between all three patient groups. For reference, individual patient data from the CVD-free patient cohort is shown (**Figure 1K-O**) with patients binned into four larger age groups.

To illustrate longitudinal developmental changes in cardiac electrical activity, vectorcardiographic QRS and T loops derived from serial ECG recordings of a single CVD-free individual were examined across ages ranging from 5 months to 27 years (**Figure 2**). Across frontal, transverse, and left sagittal planes, QRS vector loops demonstrate progressive changes in both orientation and magnitude with increasing age. In early life (5 months–5 years), QRS loops are larger in magnitude and show greater spatial dispersion, whereas with advancing age the loops become more compact and exhibit a gradual reduction in vector magnitude. Similar age-related changes are observed consistently across all three orthogonal views, indicating coordinated maturation of ventricular depolarization geometry over time. T vector loops show comparatively less variability in orientation across age, but also demonstrate a gradual reduction in magnitude with increasing age. Together, these longitudinal observations highlight age-dependent adaptations in both depolarization and repolarization vectors within the same individual. Because of the unavailability of sufficient longitudinal replicates, we were unable to repeat this analysis in a larger cohort.

**Figure 2:**
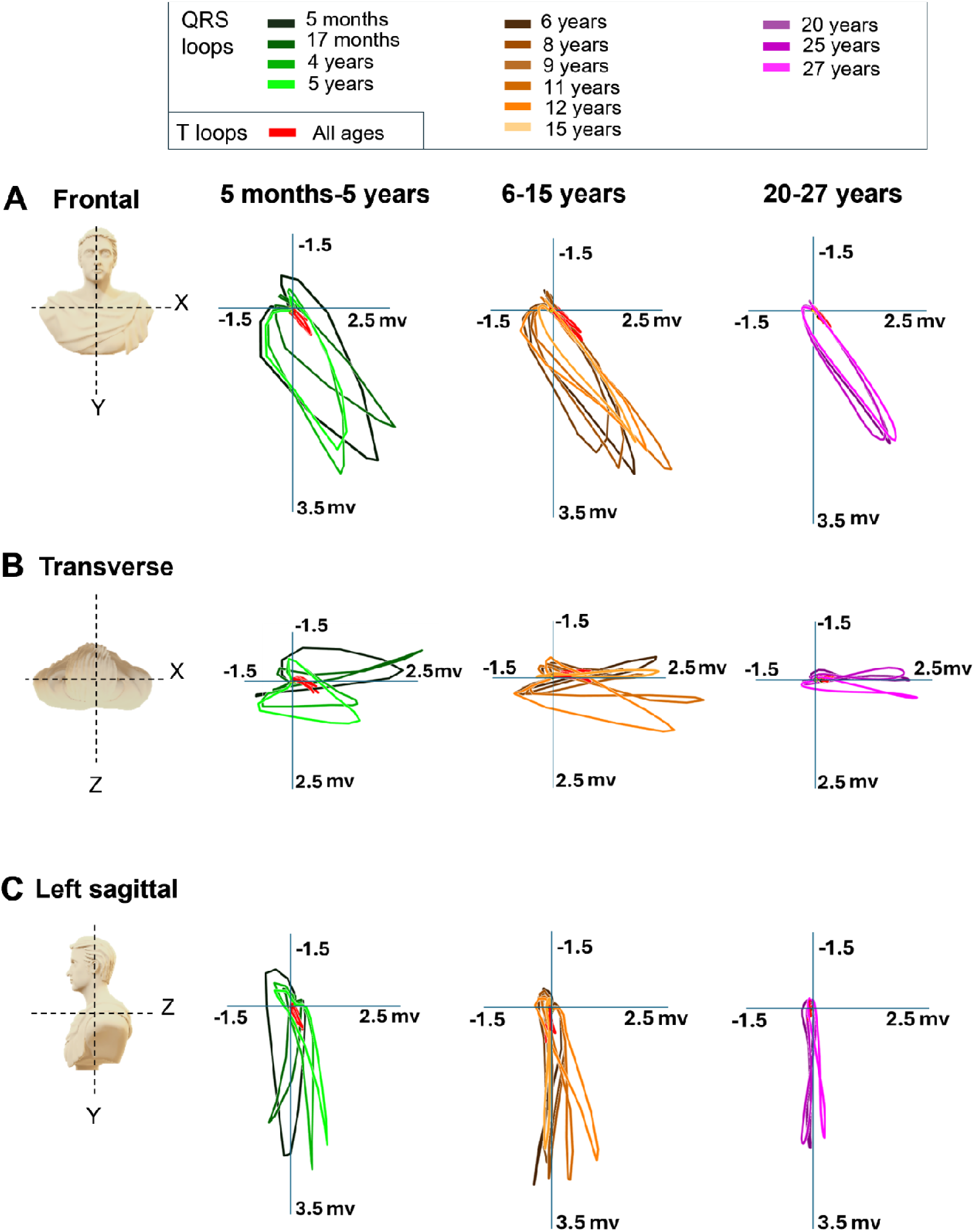
Example of developmental adaptations in longitudinal vectorcardiographic loops. Vectorcardiographic QRS and T loops were derived from serial 12-lead ECG recordings collected longitudinally from a single CVD-free female patient between the ages of 5 months and 27 years. Loops are displayed in **(A)** frontal, **(B)** transverse, and **(C)** left sagittal planes. The X lead represents the left-to-right axis, the Y lead represents the head-to-foot axis, and the Z lead represents the front-to-back axis. Patient age at the time of ECG acquisition is denoted by color, illustrating age-related changes in loop orientation and magnitude over time. This longitudinal example visually demonstrates developmental adaptations in ventricular depolarization (QRS) and repolarization (T) vectors across childhood into adulthood.

### Comparison of age-dependent changes in ECG metrics with published datasets

We compared the ECG values derived from CVD-free pediatric patients at Children’s National Hospital with ECG reference standards previously reported for healthy children. For this comparison, ECG datasets from the Pediatric Heart Network and Bratincsák, et al. studies were binned into four comparable age groups. Similar ECG values were reported across all three studies (**Figure 3A-E**). There was no statistical difference in ECG values between the current study and PHN dataset. Our age-dependent results were also similar to the Bratincsák study, although a direct statistical comparison was not feasible (raw data was not reported and Z-scores were adjusted to body surface area).

**Figure 3:**
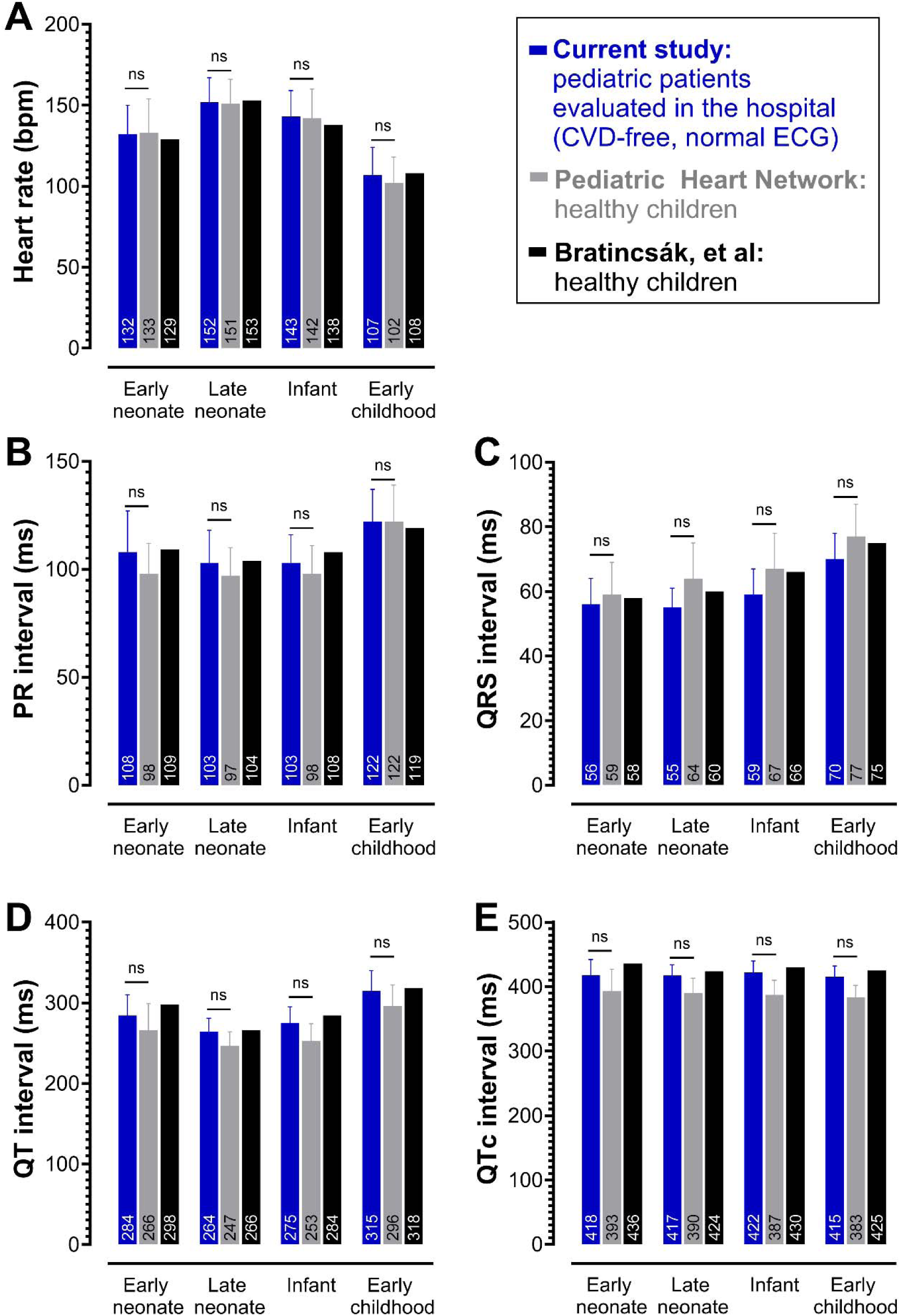
Comparison of pediatric ECG parameters between studies. ECG datasets from the CVD-free patient cohort, Pediatric Heart Network, and Bratincsák study were binned into four comparable age groups. Mean + SEM values are reported for **A)** heart rate, **B)** PR interval, **C)** QRS interval, **D)** QT interval, and **E)** Bazett corrected QT interval (note: SEM not available for Bratincsák study). Mean values are reported at the bottom of each bar plot. Unpaired two-tailed T-test was used to compare values between the current study and the pediatric heart network study; ns = not significant.

**Figure 4.**
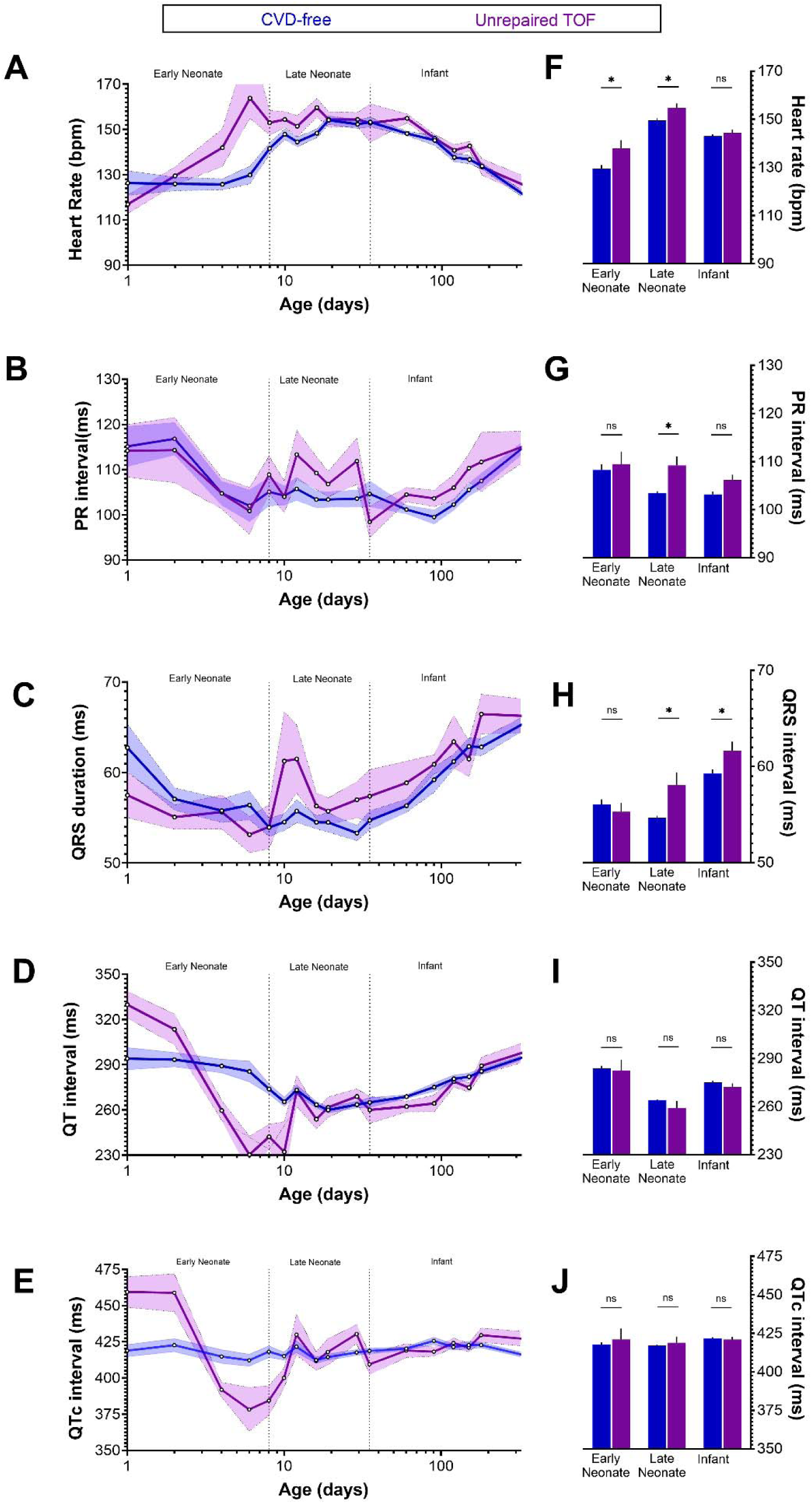
Age-dependent variations in ECG metrics in patients with unrepaired tetralogy of Fallot (TOF). Derived ECG values for heart rate, PR interval, QRS duration, QT interval, and QTc interval are shown. **A-E)** Mean values for pediatric patients with normal ECG recordings, reported across 17 age groups from neonatal to infancy. Data shown for age-matched CVD-free patients (blue) and TOF diagnosis (purple). **F-J)** Statistical comparisons between CVD-free and TOF patients, binned into three age groups. Data displayed as mean + SEM; Unpaired two-tailed T-test, *p<0.05, ns = not significant.

### Case-study in pediatric patients with tetralogy of Fallot

We examined whether developmental trends in ECG metrics might differ in patients with congenital heart disease (**Supplemental Table 3**). As a case study, we focused on pediatric patients with unrepaired TOF, a common congenital heart defect that is often associated with cyanosis^25^. Slightly faster heart rates were recorded in TOF patients during both the early neonatal (TOF: 137.9±3.4 bpm; CVD-free: 129.5±1.5 bpm) and late neonatal stages (TOF: 154.9±1.7 bpm; CVD-free: 149.5±0.9 bpm; **Figure 5**). TOF patients also displayed a longer PR interval in the late neonatal stage (TOF: 109.2±1.9 ms; CVD-free: 103.4±0.4 ms) and a slightly longer QRS in the late neonatal (TOF: 58.1±1.3 ms; CVD-free: 54.7±0.2 ms) and infant stages (TOF: 61.7±0.9 ms; CVD-free: 59.3±0.4 ms). No significant difference was detected in the QT or QTc intervals; however, larger day-to-day variations were observed in the TOF patients, particularly in the early neonatal stage.

**Figure 5.**
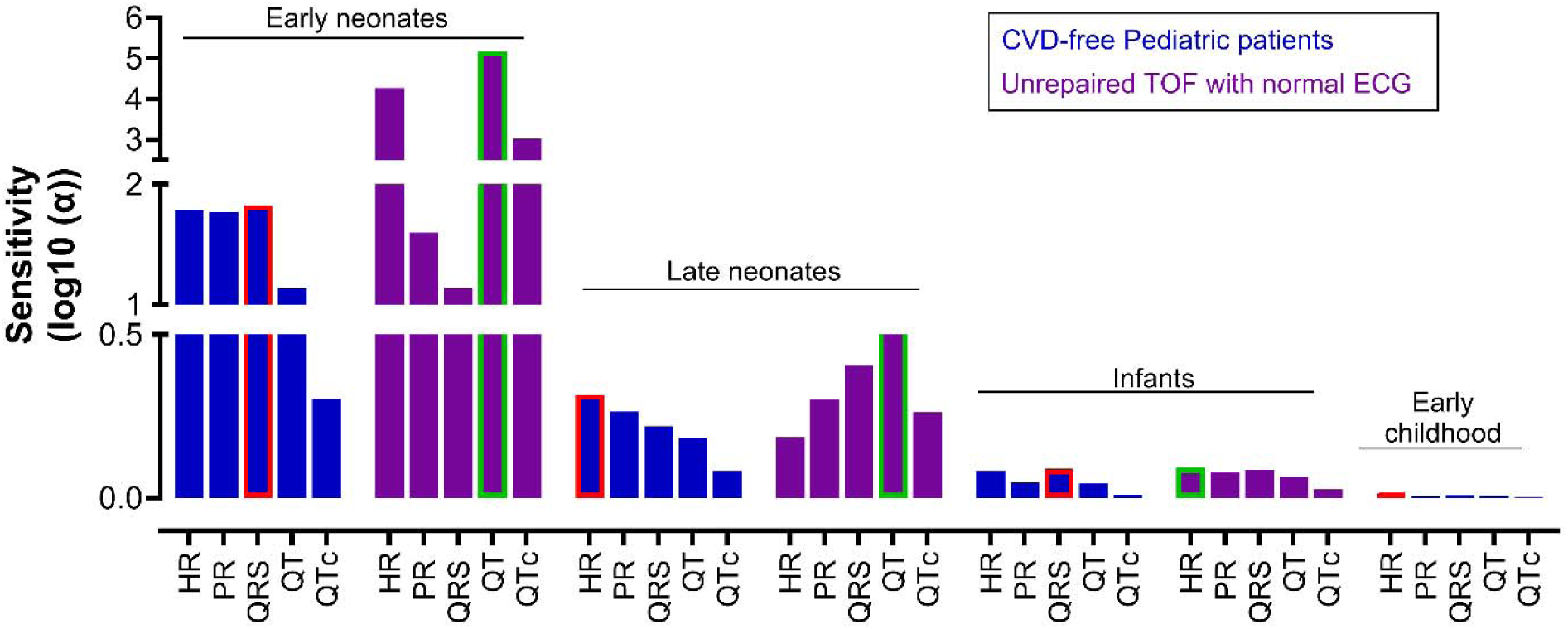
Age-dependent ECG sensitivity analysis. Sensitivity analysis of mean ECG values in CVD-free pediatric patients (blue) compared to those with unrepaired tetralogy of Fallot (purple). The most sensitive metric is highlighted for each age and patient group, which reflects the largest percent change per day of life. The sensitivity parameter “α” is defined as the % change/day = (age span)x(median value)x(95^th^ percentile – 5^th^ percentile)x100. Age span = (age end – age start) for each age group.

Next, we performed sensitivity analysis to identify the most sensitive ECG metric per age group, which reflects the percent change per day of life (**Figure 5**). As expected, the largest day-to-day changes in ECG metrics were observed in early neonates in both CVD-free and TOF patient cohorts. In CVD-free early neonates, the QRS > HR > PR values were highly sensitive to patient age (α=59-68%). In TOF early neonates, the QT > HR > QTc values were highly sensitive to patient age, displaying more exaggerated day-to-day variations (α=1,059-15,000%). More modest daily ECG changes were observed in older neonates. In CVD-free late neonates, HR > PR > QRS values were sensitive to patient age (α=1.7-2%). While in TOF late neonates, QT > QRS > PR values were sensitive to patient age (α=2-3.5%). In infants, the most sensitive ECG metric was the QRS interval in CVD-free patients (α=1.2%) and HR in TOF patients (α=1.2%). In the young childhood group, HR (α=1%) was the most sensitive ECG metric for the CVD-free patient cohort. Analysis was not performed on TOF patients for this older age group, as patients had largely undergone surgical repair by this timepoint. The results suggest that disease impacts the developmental trajectory of ECG parameters.

Finally, we derived Z-scores from the baseline CVD-free patient cohort and measured disease-specific deviations in the unrepaired TOF cohort – defined as ≥ 2 SD of baseline (e.g., HR early neonates: ±35.9 bpm, late neonates: ±29.78 bpm, infant: ±31.6bpm; **Figure 6**). Preoperative TOF early neonates had the greatest percentage of patients with ECG outlier values beyond the Z-score range (HR: 13.5%, PR: 3.8%, QRS: 3.8%, QT: 25%, QTc: 32.7% of patients), followed by late neonates (HR: 12.2%, PR: 5.5%, QRS: 6.8%, QT: 23%, QTc: 24.3%), and then infants (HR: 2.5%, PR: 8%, QRS: 5%, QT: 9.3%, QTc: 17.3%).

**Figure 6.**
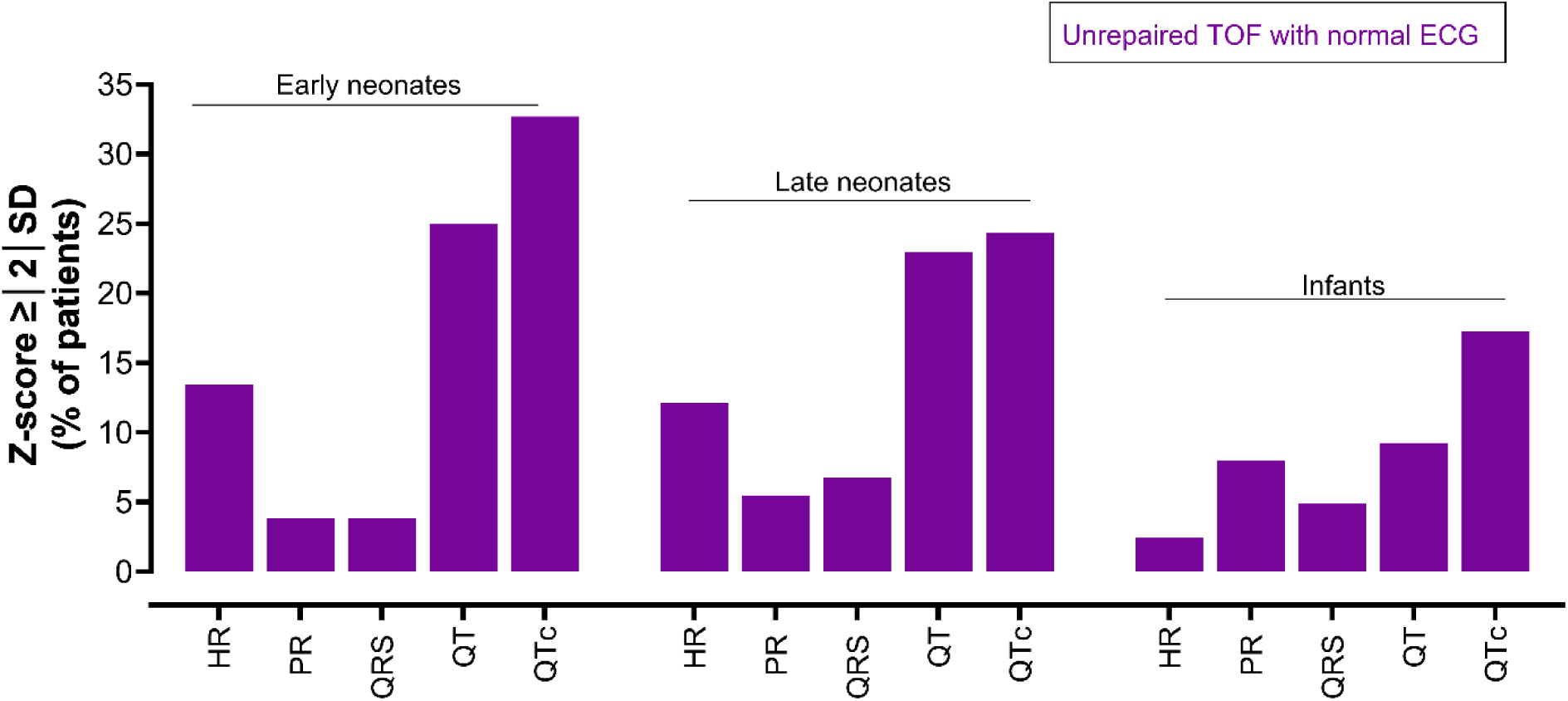
Distribution of ECG outlier values across age groups in unrepaired tetralogy of Fallot patients. Percentage of TOF patients with ECG outlier values – identified outside the range of Z-scores by ≥ |2| standard deviation; Z-score determined by baseline “normal” values from CVD-free patients. Data shown for heart rate (HR), PR Interval, QRS Complex, QT Interval, and QTc interval.

## DISCUSSION

In this study, we characterized high–temporal-resolution developmental trajectories for five clinically important ECG parameters (HR, PR interval, QRS duration, QT interval, and QTc) across multiple pediatric cohorts whose ECGs were reviewed and adjudicated as normal by a cardiologist. First, we identified age-dependent variability in key ECG parameters, underscoring the importance of studies focused on developmental cardiac electrophysiology. Second, we observed no significant differences in the developmental trajectories of ECG parameters across pediatric cohorts (all patients, ICU-excluded, and CVD-free). Third, ECG parameters in the CVD-free cohort were not significantly different from those reported in healthy children in prior studies, suggesting that developmental ECG trajectories derived from pediatric patients without cardiovascular disease accurately reflect baseline values—particularly in younger neonatal age groups where data from healthy individuals are limited. Fourth, sensitivity analyses identified ECG parameters with heightened developmental relevance at specific life stages (e.g., QRS duration in early neonates and HR in late neonates), and demonstrated that these normal developmental signatures are altered in diseased cohorts (e.g., unrepaired TOF, with QT affected in both early and late neonates). Finally, we show that high–temporal-resolution ECG trajectories in CVD-free pediatric patients provide a robust reference framework for identifying ECG outlier values, as illustrated in patients with unrepaired TOF.

### High-temporal-resolution ECG parameter developmental trajectory

In this study, we report - for the first time, high-temporal-resolution baseline reference values for five clinically important ECG parameters with day-level resolution for neonates and month-level resolution for infants. Prior studies have reported normative ECG values across age groups ranging from the neonatal period through young adulthood^5–10^, however, temporal resolution across studies was inconsistent. While some studies applied finer age stratification during the neonatal and infancy periods, age groupings typically broadened in older children, and even early-life stratification relied on discrete age categories that smooth over continuous developmental change. In contrast, our approach introduces day-level resolution in neonates and month-level resolution in infancy, enabling detection of age-group variability that is not observable when data are aggregated into broad age bins. Thus, this work extends prior age-stratified frameworks by transitioning from discrete age categories to continuous, high–temporal-resolution developmental trajectories.

### Developmental trends in ECG parameters remain conserved across patient cohorts

In the current study, we did not find significant differences in ECG parameters across pediatric patient cohorts, suggesting that overall ECG maturation is largely preserved despite the presence of diverse conditions. This finding is particularly important given that pediatric cardiovascular research frequently relies on retrospective clinical datasets rather than prospective trials. For example, a recent study constructed a pediatric ECG database using retrospective data from more than 11,000 patients with a wide range of CVD diagnoses^26^, while Bratincsák et al. ^5^ retrospectively analyzed ECGs from over 27,000 children and young adults to establish standardized normative Z-score values. Despite the widespread use of such datasets, it has remained unclear whether normal developmental ECG trajectories are maintained in retrospectively collected clinical cohorts. Although non-CVD illnesses could plausibly influence cardiovascular development and ECG parameters, our findings indicate that these effects are not significant at the population level. Importantly, this does not preclude the possibility that specific CVD diagnoses may alter developmental ECG trajectories (our dataset only included patients whose ECGs were reviewed and adjudicated as normal by a cardiologist.); however, as demonstrated here, such condition-specific effects are not apparent when data are analyzed at the aggregated cohort level.

### ECG parameters from pediatric CVD-free cohort may serve as baseline

In this study, we found that ECG parameters in a CVD-free pediatric cohort do not differ significantly from those reported in healthy pediatric populations, as described by Bratincsák et al. ^5^ and the PHN ECG study ^8^. These findings suggest that retrospectively derived ECG values from CVD-free pediatric patients can serve as reliable baseline reference data. Prospective enrollment of pediatric subjects is often limited by small patient numbers, ethical considerations, and the need for parental consent^27,28^. In addition, identifying truly healthy children for ECG studies is challenging due to the extensive review and adjudication required in large clinical datasets. As a result, retrospectively derived ECG parameters from CVD-free patients that align with those from healthy cohorts offer a practical alternative, reducing time and resource demands while avoiding key limitations of prospective studies. Preservation of normal developmental ECG trajectories in CVD-free patients, comparable to those observed in healthy pediatric populations, is therefore not unexpected. Although developmental delays have been reported in children with congenital heart disease^29–31^, there is no clear evidence that non-CVD conditions independently alter cardiac electrophysiological maturation.

### Unrepaired TOF alters developmental ECG trajectories

Analysis of the unrepaired TOF cohort revealed significant deviations in ECG parameters relative to CVD-free reference values, indicating disease-related alterations in developmental ECG trajectories. These deviations were age dependent and non-linear, with distinct patterns across ECG parameters. For example, heart rate was higher in unrepaired TOF patients during early and late neonatal periods, whereas QRS duration was prolonged in late neonates and infants compared with the CVD-free cohort (**Figure 4**). Sensitivity analyses further demonstrated age-dependent increases in parameter sensitivity in unrepaired TOF patients. Notably, the QT interval emerged as the most age-sensitive parameter in early and late neonates, and Z-score analysis identified QT and QTc as having the highest frequency of outliers in these same age groups (**Figure 5**). Collectively, these findings indicate that unrepaired TOF disrupts ECG development in an age- and parameter-specific manner, with repolarization measures showing particular vulnerability early in life.

Although characteristic ECG abnormalities in TOF—such as QRS prolongation following surgical repair and features of right ventricular hypertrophy—are well described^32–35^, these observations largely derive from post-repair cohorts. To our knowledge, few studies have directly compared ECG metrics in unrepaired TOF patients with control populations, as prior work has primarily focused on associations between ECG abnormalities and adverse outcomes after repair ^32,33,36^. In this context, our Z-score–based identification of abnormal ECG values in unrepaired TOF patients highlights a potential clinical application of developmental ECG reference trajectories for detecting early electrophysiological deviations that may inform risk stratification before surgical intervention.

### Limitations

Several limitations should be considered when interpreting these findings. First, analyses were based on retrospectively collected clinical ECG data, limiting control over acquisition conditions and ECG system variability. While retrospective datasets enable large sample sizes and reflect real-world clinical practice, they do not permit causal inference. Second, a sufficient number of samples (e.g., n=33-237 per age unit in “all patients” cohort) were included for statistical analysis between age groups; however, larger sample sizes could refine age-specific means and potentially shift observed developmental trends. Third, the cohort was hospital based rather than population based. Although developmental ECG trajectories were preserved across in-hospital cohorts, subtle influences of non-cardiac comorbidities or clinical context cannot be fully excluded. Fourth, developmental trajectories were derived from cross-sectional, age-stratified data rather than longitudinal follow-up, precluding direct assessment of within-subject changes over time. To partially address this limitation, longitudinal ECGs from a CVD-free individual demonstrated expected developmental changes in vectorcardiographic QRS and T loops (**Figure 2**). Fifth, the unrepaired TOF cohort likely represents a heterogeneous population with variable disease severity, which was not explicitly stratified and may contribute to ECG variability beyond age-related effects. Finally, analyses were limited to standard clinically reported ECG parameters; additional electrophysiological markers, such as measures of repolarization heterogeneity or conduction dispersion, were not assessed and warrant investigation in future studies.

## Conclusions

In conclusion, we report high–temporal-resolution reference values for five clinically important ECG parameters, with day-level resolution in neonates and month-level resolution in infants.

Our findings demonstrate that retrospectively collected ECG data from a CVD-free pediatric cohort closely mirror values reported in healthy pediatric populations, supporting their use as surrogate baseline reference data. Finally, while developmental ECG trajectories are preserved in CVD-free cohorts, they diverge in disease states such as unrepaired TOF, highlighting the potential clinical utility of these reference trajectories for identifying disease-related alterations in developmental cardiac electrophysiology.

## Data Availability

All data produced in the present work are contained in the manuscript.

## ACKNOWLEDGEMENTS

We thank Bao Nguyen Puente from the Cardiac Critical Care Unit for valuable scientific insight that informed cohort classification and supported accurate interpretation of the data. We also acknowledge the NIH/NHLBI Pediatric Heart Network Echo-Z/Normal ECG dataset, which was used in preparation of this work. Data was downloaded from www.pediatricheartnetwork.org on 02/11/2025.

## Supplemental Materials

Supplemental Tables 1-3

