## Supplemental Table 1 for "Age-dependent Dynamics of the Electrocardiographic Parameters in Cardiovascular Disease-Free Children"

| Age (days) | HR (mean) | HR (SEM) | PR (mean) | PR (SEM) | QRS (mean) | QRS (SEM) | QT (mean) | QT (SEM) | QTc (mean) | QTc (SEM) |
| --- | --- | --- | --- | --- | --- | --- | --- | --- | --- | --- |
| 1 | 126.49 | 3.86 | 117.41 | 3.73 | 63.03 | 1.97 | 296.86 | 6.30 | 424.19 | 6.29 |
| 2 | 129.33 | 2.30 | 116.46 | 2.91 | 56.81 | 0.91 | 288.13 | 3.46 | 420.13 | 3.19 |
| 3 | 132.32 | 1.71 | 109.10 | 2.48 | 55.73 | 1.23 | 289.06 | 3.84 | 425.80 | 4.40 |
| 4 | 129.64 | 2.13 | 110.24 | 2.46 | 56.83 | 0.95 | 287.01 | 3.11 | 416.83 | 2.61 |
| 5 | 132.32 | 2.40 | 105.93 | 2.15 | 54.13 | 0.87 | 285.44 | 3.43 | 420.84 | 3.86 |
| 6 | 133.91 | 2.64 | 105.03 | 3.29 | 55.76 | 1.11 | 281.21 | 5.47 | 416.58 | 6.38 |
| 7 | 142.93 | 1.63 | 109.96 | 2.16 | 53.43 | 1.24 | 271.13 | 2.25 | 415.42 | 2.51 |
| 8 | 142.77 | 1.96 | 104.28 | 1.84 | 55.18 | 0.73 | 268.46 | 2.98 | 410.53 | 3.05 |
| 9 | 147.92 | 1.58 | 105.28 | 1.65 | 54.86 | 0.61 | 261.51 | 1.82 | 408.67 | 1.93 |
| 10 | 149.55 | 1.50 | 104.82 | 1.53 | 55.15 | 0.73 | 263.87 | 1.88 | 414.11 | 2.28 |
| 11 | 149.65 | 1.64 | 105.57 | 1.94 | 55.70 | 0.68 | 264.70 | 2.10 | 415.76 | 2.40 |
| 12 | 148.28 | 1.48 | 106.28 | 1.61 | 55.06 | 0.80 | 267.49 | 1.88 | 418.14 | 2.37 |
| 13 | 151.71 | 1.60 | 106.08 | 1.60 | 56.02 | 0.64 | 262.45 | 1.85 | 414.96 | 1.96 |
| 14 | 144.46 | 1.48 | 104.27 | 1.62 | 56.04 | 0.62 | 270.16 | 1.91 | 416.34 | 1.94 |
| 15 | 150.67 | 1.42 | 105.27 | 1.36 | 56.06 | 0.71 | 262.95 | 1.82 | 414.35 | 1.95 |
| 16 | 148.39 | 1.36 | 103.27 | 1.39 | 56.00 | 0.62 | 265.67 | 1.70 | 415.81 | 1.98 |
| 17 | 150.36 | 1.46 | 105.69 | 1.47 | 55.43 | 0.57 | 265.34 | 1.85 | 417.96 | 2.11 |
| 18 | 152.97 | 1.50 | 103.21 | 1.24 | 56.99 | 1.35 | 260.77 | 2.84 | 414.14 | 3.95 |
| 19 | 152.72 | 1.38 | 104.60 | 1.40 | 56.58 | 0.76 | 262.64 | 1.67 | 417.28 | 1.76 |
| 20 | 149.26 | 1.48 | 101.31 | 1.14 | 56.00 | 0.60 | 268.09 | 2.27 | 419.81 | 2.47 |
| 21 | 153.22 | 1.02 | 104.30 | 1.12 | 55.34 | 0.50 | 262.68 | 1.28 | 417.92 | 1.64 |
| 22 | 155.12 | 1.23 | 103.67 | 1.31 | 54.70 | 0.56 | 259.56 | 1.52 | 415.21 | 1.87 |
| 23 | 155.81 | 1.25 | 102.29 | 1.39 | 54.87 | 0.56 | 260.77 | 1.57 | 418.21 | 1.53 |
| 24 | 151.23 | 1.43 | 104.67 | 1.32 | 55.40 | 0.61 | 264.76 | 1.99 | 419.27 | 2.12 |
| 25 | 153.23 | 1.70 | 105.51 | 1.48 | 54.70 | 0.56 | 263.90 | 1.85 | 419.60 | 1.67 |
| 26 | 152.10 | 1.58 | 102.37 | 1.39 | 55.07 | 0.71 | 265.09 | 1.92 | 420.57 | 1.97 |
| 27 | 151.89 | 1.38 | 105.90 | 1.22 | 55.65 | 0.57 | 266.62 | 1.48 | 422.62 | 1.71 |
| 28 | 152.49 | 1.44 | 103.46 | 1.39 | 55.74 | 0.58 | 266.43 | 2.07 | 422.32 | 2.09 |
| 29 | 154.12 | 1.09 | 102.79 | 1.17 | 54.43 | 0.55 | 262.30 | 2.09 | 420.14 | 1.36 |
| 30 | 154.70 | 1.23 | 101.48 | 1.00 | 55.10 | 0.60 | 264.65 | 1.49 | 422.82 | 1.48 |
| 31 | 154.52 | 1.47 | 102.30 | 1.36 | 54.83 | 0.77 | 262.34 | 2.17 | 419.06 | 2.91 |
| 32 | 157.91 | 1.27 | 104.80 | 1.89 | 54.71 | 0.67 | 260.71 | 1.63 | 421.02 | 1.70 |
| 33 | 153.48 | 1.51 | 102.78 | 1.61 | 54.98 | 1.00 | 263.82 | 1.78 | 420.48 | 2.28 |
| 34 | 152.78 | 1.62 | 104.78 | 1.58 | 55.22 | 0.66 | 264.09 | 1.79 | 419.09 | 1.94 |
| 35 | 154.61 | 1.28 | 103.16 | 1.80 | 55.21 | 0.78 | 262.71 | 1.90 | 418.81 | 1.73 |
| 60 | 147.01 | 0.80 | 101.46 | 0.73 | 57.19 | 0.56 | 270.01 | 0.99 | 420.31 | 0.95 |
| 90 | 141.34 | 1.63 | 99.87 | 1.28 | 60.90 | 0.90 | 278.68 | 1.81 | 424.63 | 1.57 |
| 120 | 139.40 | 1.45 | 102.44 | 1.03 | 61.38 | 0.59 | 280.23 | 1.86 | 423.86 | 1.64 |
| 150 | 138.64 | 1.36 | 104.04 | 0.98 | 62.33 | 0.62 | 280.21 | 1.62 | 422.88 | 1.58 |
| 180 | 133.19 | 1.01 | 106.71 | 1.11 | 63.18 | 0.63 | 285.25 | 1.31 | 421.95 | 1.49 |
| 365 | 118.39 | 1.56 | 116.87 | 0.96 | 65.87 | 0.52 | 295.91 | 1.32 | 416.01 | 1.22 |
| 730 | 114.26 | 1.01 | 119.64 | 0.91 | 67.12 | 0.52 | 299.05 | 1.28 | 407.65 | 0.85 |
| 1095 | 105.98 | 1.17 | 123.85 | 1.11 | 71.14 | 0.56 | 320.34 | 1.71 | 420.38 | 1.31 |
| 1460 | 101.96 | 1.18 | 122.42 | 1.12 | 73.70 | 0.61 | 326.21 | 1.57 | 419.99 | 1.34 |
| 1825 | 96.17 | 1.28 | 127.65 | 1.27 | 76.08 | 0.70 | 335.37 | 1.85 | 419.63 | 1.34 |

**Table 1. ECG values for non-ICU cohort (n=5,007).**
