## Supplemental Table 2 for "Age-dependent Dynamics of the Electrocardiographic Parameters in Cardiovascular Disease-Free Children"

| Age (days) | HR (mean) | HR (SEM) | PR (mean) | PR (SEM) | QRS (mean) | QRS (SEM) | QT (mean) | QT (SEM) | QTc (mean) | QTc (SEM) |
| --- | --- | --- | --- | --- | --- | --- | --- | --- | --- | --- |
| 1 | 126.29 | 5.44 | 115.14 | 4.46 | 62.76 | 2.62 | 294.00 | 7.36 | 419.10 | 4.32 |
| 2 | 125.92 | 3.04 | 116.81 | 3.61 | 57.08 | 1.24 | 293.39 | 4.76 | 423.08 | 4.60 |
| 3 | 130.83 | 2.38 | 106.46 | 3.06 | 55.83 | 1.61 | 285.15 | 3.49 | 420.13 | 3.89 |
| 4 | 125.74 | 2.48 | 104.76 | 2.67 | 55.79 | 1.31 | 289.14 | 4.50 | 414.52 | 4.70 |
| 5 | 127.88 | 3.95 | 107.92 | 3.81 | 53.46 | 1.25 | 289.22 | 4.70 | 418.96 | 4.29 |
| 6 | 129.87 | 3.95 | 102.00 | 3.53 | 56.40 | 1.61 | 285.47 | 6.95 | 416.47 | 5.69 |
| 7 | 142.19 | 1.85 | 107.33 | 2.92 | 53.26 | 1.81 | 269.16 | 2.35 | 412.40 | 2.81 |
| 8 | 141.59 | 2.59 | 105.09 | 3.17 | 53.94 | 1.01 | 273.88 | 3.41 | 418.15 | 4.01 |
| 9 | 150.32 | 2.75 | 102.54 | 2.28 | 53.35 | 0.90 | 260.11 | 2.90 | 409.30 | 2.55 |
| 10 | 147.88 | 2.44 | 104.24 | 2.16 | 54.54 | 0.94 | 265.41 | 3.06 | 414.12 | 2.55 |
| 11 | 148.26 | 2.54 | 103.79 | 2.91 | 56.00 | 1.23 | 268.82 | 3.05 | 419.90 | 2.64 |
| 12 | 144.46 | 2.22 | 105.73 | 2.62 | 55.74 | 1.27 | 273.22 | 3.36 | 421.61 | 4.16 |
| 13 | 150.57 | 2.62 | 104.86 | 2.39 | 54.88 | 1.00 | 264.11 | 2.50 | 416.63 | 2.54 |
| 14 | 144.19 | 2.26 | 101.35 | 2.41 | 55.81 | 0.93 | 273.08 | 2.84 | 420.75 | 3.43 |
| 15 | 152.62 | 1.99 | 103.51 | 1.64 | 55.56 | 0.98 | 262.71 | 2.44 | 417.18 | 3.05 |
| 16 | 148.33 | 1.96 | 103.40 | 1.91 | 54.51 | 0.73 | 263.41 | 2.09 | 412.25 | 2.22 |
| 17 | 145.70 | 1.94 | 106.78 | 1.70 | 55.57 | 0.75 | 267.13 | 2.44 | 414.55 | 2.41 |
| 18 | 152.31 | 2.16 | 103.33 | 1.86 | 56.90 | 1.11 | 265.81 | 2.79 | 421.48 | 3.18 |
| 19 | 154.09 | 1.93 | 103.40 | 1.75 | 54.50 | 1.08 | 259.95 | 1.95 | 414.36 | 2.14 |
| 20 | 151.52 | 2.01 | 99.77 | 1.67 | 54.44 | 0.72 | 263.93 | 2.44 | 416.04 | 2.09 |
| 21 | 153.11 | 1.68 | 103.22 | 2.16 | 53.48 | 0.57 | 262.00 | 1.77 | 416.81 | 1.68 |
| 22 | 156.12 | 1.61 | 102.80 | 2.03 | 54.57 | 0.83 | 260.12 | 1.82 | 417.75 | 2.15 |
| 23 | 157.05 | 2.52 | 99.49 | 2.54 | 53.40 | 0.71 | 260.37 | 2.45 | 418.44 | 1.74 |
| 24 | 152.05 | 2.04 | 103.58 | 2.07 | 54.77 | 0.90 | 261.17 | 2.18 | 415.38 | 2.42 |
| 25 | 155.08 | 2.75 | 104.42 | 2.70 | 54.97 | 0.88 | 258.87 | 2.21 | 414.08 | 2.07 |
| 26 | 150.63 | 2.63 | 102.72 | 2.56 | 52.79 | 0.85 | 263.68 | 2.82 | 415.45 | 2.54 |
| 27 | 150.22 | 2.36 | 105.81 | 2.06 | 54.78 | 0.87 | 267.20 | 2.38 | 420.71 | 2.53 |
| 28 | 152.33 | 2.44 | 102.45 | 2.27 | 55.91 | 0.84 | 265.51 | 3.00 | 420.04 | 2.37 |
| 29 | 152.31 | 1.97 | 103.60 | 1.95 | 53.31 | 0.85 | 263.53 | 2.16 | 417.64 | 2.00 |
| 30 | 153.04 | 2.18 | 100.48 | 1.73 | 55.49 | 0.88 | 265.80 | 2.59 | 421.51 | 2.01 |
| 31 | 154.60 | 1.98 | 100.43 | 1.24 | 52.86 | 0.82 | 260.62 | 2.23 | 417.74 | 2.26 |
| 32 | 157.83 | 1.85 | 107.54 | 3.71 | 54.19 | 1.01 | 260.67 | 2.37 | 420.79 | 2.64 |
| 33 | 153.86 | 2.51 | 104.55 | 2.61 | 54.31 | 1.33 | 261.03 | 2.97 | 416.45 | 3.17 |
| 34 | 152.38 | 2.33 | 106.80 | 2.72 | 55.60 | 0.77 | 262.57 | 2.24 | 416.36 | 2.17 |
| 35 | 153.26 | 2.15 | 104.65 | 2.88 | 54.72 | 1.04 | 265.02 | 3.28 | 418.67 | 1.95 |
| 60 | 148.18 | 1.01 | 101.16 | 0.99 | 56.33 | 0.73 | 268.85 | 1.25 | 420.30 | 1.21 |
| 90 | 145.21 | 2.32 | 99.51 | 1.62 | 59.20 | 1.40 | 275.17 | 2.77 | 425.57 | 2.52 |
| 120 | 137.64 | 1.98 | 102.26 | 1.31 | 61.22 | 0.81 | 280.61 | 2.58 | 422.06 | 2.02 |
| 150 | 136.69 | 1.90 | 105.55 | 1.46 | 62.89 | 0.95 | 282.08 | 2.36 | 422.69 | 2.59 |
| 180 | 133.83 | 1.46 | 107.49 | 1.73 | 62.82 | 0.96 | 285.63 | 1.86 | 423.61 | 2.03 |
| 365 | 119.07 | 1.40 | 116.03 | 1.28 | 65.80 | 0.76 | 296.38 | 1.41 | 415.13 | 1.61 |
| 730 | 112.73 | 1.29 | 119.74 | 1.16 | 66.38 | 0.57 | 300.44 | 1.63 | 406.27 | 1.02 |
| 1095 | 106.02 | 1.38 | 124.18 | 1.33 | 70.53 | 0.61 | 319.28 | 2.02 | 419.44 | 1.50 |
| 1460 | 102.11 | 1.51 | 123.26 | 1.33 | 73.48 | 0.72 | 326.04 | 1.89 | 420.13 | 1.66 |
| 1825 | 93.79 | 1.51 | 128.50 | 1.53 | 75.75 | 0.83 | 337.20 | 2.09 | 417.53 | 1.61 |

**Table 2. ECG values for CVD-free (non-ICU, non-CVD) cohort (n=2,449).**
