## Supplemental Table 3 for "Age-dependent Dynamics of the Electrocardiographic Parameters in Cardiovascular Disease-Free Children"

| Age (days) | HR (mean) | HR (SEM) | PR (mean) | PR (SEM) | QRS (mean) | QRS (SEM) | QT (mean) | QT (SEM) | QTc (mean) | QTc (SEM) |
| --- | --- | --- | --- | --- | --- | --- | --- | --- | --- | --- |
| 1 | 116.92 | 4.11 | 114.17 | 5.82 | 57.50 | 2.49 | 329.83 | 8.93 | 459.42 | 10.58 |
| 2 | 129.46 | 4.01 | 114.31 | 7.23 | 55.08 | 1.35 | 313.46 | 10.24 | 458.85 | 13.14 |
| 4 | 141.91 | 8.31 | 104.73 | 3.46 | 55.64 | 1.88 | 259.64 | 6.68 | 391.91 | 5.14 |
| 6 | 163.86 | 8.95 | 100.86 | 5.20 | 53.14 | 1.99 | 230.29 | 12.33 | 378.29 | 15.14 |
| 8 | 153.00 | 5.44 | 108.89 | 4.14 | 54.00 | 2.45 | 242.22 | 8.37 | 384.33 | 10.28 |
| 10 | 154.36 | 3.45 | 104.00 | 3.57 | 61.27 | 5.46 | 232.18 | 20.03 | 400.00 | 6.86 |
| 12 | 151.42 | 4.71 | 113.33 | 5.38 | 61.50 | 3.77 | 272.67 | 10.38 | 429.92 | 14.18 |
| 16 | 159.62 | 4.10 | 109.23 | 3.65 | 56.31 | 1.45 | 253.85 | 6.41 | 411.69 | 6.35 |
| 19 | 154.40 | 3.48 | 106.80 | 2.82 | 55.73 | 1.48 | 261.73 | 6.81 | 417.93 | 9.11 |
| 29 | 154.38 | 3.14 | 111.88 | 5.12 | 57.00 | 1.99 | 268.88 | 5.13 | 430.38 | 6.62 |
| 35 | 152.80 | 8.55 | 98.44 | 3.51 | 57.40 | 2.94 | 260.00 | 9.02 | 409.40 | 7.00 |
| 60 | 154.91 | 1.91 | 104.48 | 1.58 | 58.87 | 2.40 | 262.22 | 3.59 | 418.91 | 4.83 |
| 90 | 146.06 | 1.66 | 103.64 | 1.83 | 60.92 | 1.05 | 264.32 | 5.57 | 418.16 | 3.18 |
| 120 | 140.76 | 2.56 | 106.00 | 2.90 | 63.42 | 2.89 | 278.87 | 3.70 | 424.13 | 3.50 |
| 150 | 142.67 | 2.08 | 110.33 | 3.11 | 61.50 | 1.97 | 274.83 | 3.05 | 421.00 | 2.89 |
| 180 | 133.38 | 3.59 | 111.69 | 6.55 | 66.46 | 2.20 | 289.23 | 5.80 | 429.62 | 4.91 |
| 365 | 124.18 | 4.03 | 115.65 | 2.93 | 66.24 | 1.82 | 299.53 | 6.62 | 426.71 | 5.15 |

**Table 3. ECG values for TOF cohort (n=305).**
